# Seroprevalence and correlates of SARS-CoV-2 neutralizing antibodies: Results from a population-based study in Bonn, Germany

**DOI:** 10.1101/2020.08.24.20181206

**Authors:** N. Ahmad Aziz, Victor M. Corman, Antje K.C. Echterhoff, Anja Richter, Antonio Schmandke, Marie Luisa Schmidt, Thomas H. Schmidt, Folgerdiena M. de Vries, Christian Drosten, Monique M.B. Breteler

## Abstract

**Background:** Accurate estimates of SARS-CoV-2 seroprevalence are crucial for the implementation of effective public health measures, but are currently largely lacking in regions with low infection rates. This is further complicated by inadequate test performance of many widely used serological assays. We therefore aimed to assess SARS-CoV-2 seroprevalence in a region with low COVID-19 burden, especially focusing on neutralizing antibodies that presumably constitute a major component of acquired immunity.

**Methods:** We invited all individuals who were enrolled in the Rhineland Study, an ongoing community-based prospective cohort study in people aged 30 years and above in the city of Bonn, Germany (N=5427). Between April 24^th^ and June 30^th^, 2020, 4771 (88%) of these individuals participated in the serosurvey. Anti-SARS-CoV-2 IgG levels were measured using an ELISA assay, and all positive or borderline results were subsequently examined through both a recombinant immunofluorescent assay and a plaque reduction neutralisation test (PRNT).

**Findings:** Seroprevalence was 0·97% (95% CI: 0·72–1·30) by ELISA and 0·36% (95% CI: 0·21–0·61) by PRNT, and did not vary with either age or sex. All PRNT+ individuals reported having experienced at least one symptom (odds ratio (OR) of PRNT+ for each additional symptom: 1·12 (95% CI: 1·04–1·21)). Apart from living in a household with a SARS-CoV-2 confirmed or suspected person, a recent history of reduced taste or smell, fever, chills/hot flashes, pain while breathing, pain in arms/legs, as well as muscle pain and weakness were significantly associated with the presence of neutralizing antibodies in those with mild to moderate infection (ORs 3·44 to 9·97, all p<0·018).

**Interpretation:** Our findings indicate a relatively low SARS-CoV-2 seroprevalence in Bonn, Germany (until June 30^th^, 2020), with neutralizing antibodies detectable in only one third of those with a positive immunoassay result, implying that almost the entire population in this region remains susceptible to SARS-CoV-2 infection.

**Funding:** The Rhineland Study is predominantly funded through the German Center for Neurodegenerative Diseases (DZNE) by the Federal Ministry of Education and Research (BMBF) and the Ministry of Culture and Science of the German State of North Rhine-Westphalia. The National Consultant Laboratory for Coronaviruses is funded by the Federal Ministry of Health (BMG). No additional funding was received for this seroprevalence study.

## INTRODUCTION

A rapidly developing global outbreak of infections by the severe acute respiratory syndrome coronavirus 2 (SARS-CoV-2) has led to the current pandemic of coronavirus disease 2019 (COVID-19).^1^ As of August 21^th^ 2020, the virus has infected more than 22·5 million people worldwide, resulting in more than 792,000 deaths.^2^ Accurate estimates of SARS-CoV-2 seroprevalence patterns in the general population are crucial for developing effective strategies to deal with the pandemic and its sequelae.^3,4^ Population studies are the only way to gain knowledge about the prevalence of asymptomatic and mildly symptomatic cases, which is of paramount importance as such individuals may elude the classical symptom-based infection chain tracking methods, but yet play a key role in the further spreading and sustainment of the current global outbreak.^5^ Moreover, seroprevalence studies provide important benchmarks for tracking the evolution of the pandemic by enabling incidence estimates at population-level.^6^

Several population-based SARS-CoV-2 serosurveillance studies have already been performed around the globe. They were predominantly conducted in areas with a disproportionately high number of COVID-19-related hospitalizations and mortality rates, with widely varying seroprevalence estimates.^7-13^ Many of these estimates may have been biased,^11^ due to inadequate sampling methods, poor antibody test performance, non-random sampling (e.g. self-referral), a non-representative sampling setting (e.g. blood donors), as well as small sample sizes.^11^ In Germany, findings of only a few small-scale serosurveys have been reported. Only two of these were population-based studies,^14,15^ while the remaining assessments targeted industrial workers, health-care providers, mothers, students/teachers or blood donors.^12^ COVID-19-related morbidity and mortality rates have been relatively low in Germany compared to other (European) countries,^16,17^ yet the true exposure state of the population could be much higher given the unknown proportion of SARS-CoV-2 infections with mild or asymptomatic course.^9^

An important challenge for accurate assessment of SARS-CoV-2 seroprevalence, especially in regions with a relatively low infection rate, is insufficient specificity of the serological tests. Widely used (point-of-care) lateral flow and quantitative ELISA assays lead to a relatively large number of false positives due to cross-reactivity with other (endemic) coronaviruses.^18^ The current gold standard for SARS-CoV-2 serology are neutralization assays.^18^ The presence of antibodies that can neutralize the virus not only is highly specific for having sustained an infection, but is also thought to constitute a major component of the acquired immunity to the virus.^19^ However, neutralization assays are highly laborious, can only be performed in specialized (i.e. biosafety level-3) laboratories, and hitherto have hardly been used in serosurveys. Thus, currently little is known about the determinants and correlates of SARS-CoV-2 neutralizing antibodies at population-level.

We aimed to *1)* accurately estimate the prevalence of SARS-CoV-2 seropositivity in a region with a relatively low infection rate, using a multi-tiered serological testing strategy that includes confirmatory neutralization assays, and *2)* investigate the correlates of neutralizing antibodies against SARS-CoV-2, with a particular focus on the characteristics of infected but asymptomatic or mildly/moderately symptomatic individuals. By embedding a large-scale seroepidemiological study within the pre-existing framework of an ongoing prospective community-based cohort study, we intended to prevent self-referral bias, ensure long-term follow-up of the participants (including future seroconversion), and facilitate the investigation of genetic, health and lifestyle determinants of susceptibility and resilience to SARS-CoV-2 infection. In this report we present the cross-sectional findings of the first serosurvey.

## METHODS

### Study population

The study was based on the Rhineland Study, an ongoing community-based cohort study in Bonn, Germany. All inhabitants aged 30–100 years of two geographically defined areas are invited to participate in the Rhineland Study. The sole exclusion criterion is insufficient command of the German language to provide informed consent. Persons living in the recruitment areas are predominantly German from Caucasian descent. The Rhineland Study’s overarching aims are to investigate the etiology and prediction of age-related (neurodegenerative) diseases and to assess normal and pathological (brain) structure and function over the adult life course.^20^ The study started in 2016 and emphasizes deep phenotyping. Because of the imposition of local lockdown measures, regular study visits were paused on March 18^th^, 2020.

This serosurvey was conducted in two groups. Group I consisted of all living participants who had been enrolled in the Rhineland Study until March 18, 2020 (N=5427). Group II consisted of individuals who were eligible for but had not yet participated in the Rhineland Study and who actively approached us to indicate their willingness to participate in the serosurvey and become prospective participants (N=597).

Approval to undertake the Rhineland Study was obtained from the ethics committee of the University of Bonn, Medical Faculty. The Rhineland Study is carried out in accordance with the recommendations of the International Conference on Harmonization (ICH) Good Clinical Practice (GCP) standards (ICH-GCP) after obtainment of written informed consent from all participants in accordance with the Declaration of Helsinki. No separate ethical approval for this serosurvey was required given its embedding in the Rhineland Study, the ethical mandate of which already covered follow-up measurements, including collection of serial bio-samples.

### Study design and procedures

All participants were informed about the serosurvey through email, postal letter and/or phone. All invitees were requested to use a purpose-designed online platform to make an appointment at one of the two local study centers, except when they were suffering from symptoms of an acute infection (especially fever, cough or other flu-like complaints), in which case they were recommended to visit a doctor. From April 24^th^ through June 30^th^, 4771 (88%) of the invited participants from Group I and 360 (60%) of the invited participants from Group II, visited one of the study centers for a blood withdrawal. Reasons for non-response included death, undeliverable invitations, and refusal due to a perceived high burden or risk of infection due to old age, immobility or co-morbidity.

### Data collection

Data collection included blood withdrawal and questionnaires. At the study center, blood was collected from an antecubital or dorsal hand vein. Thirty to 90 min after blood withdrawal serum tubes were centrifuged for 15 minutes at 2000 *g*, stored directly at +2 to +8 °C thereafter, and sent to the diagnostic lab via overnight courier within about one week of collection. Following blood collection, participants received a paper questionnaire that they were asked to complete at home and return to the study centre.

### Serological measurements

Serological analyses for SARS-CoV-2 were performed at the Institute of Virology (Charité – Universitätsmedizin Berlin, Germany) using a three-tired approach. First, the levels of IgG antibodies against SARS-CoV-2 were measured using a commercially available ELISA assay (Euroimmun, Lübeck, Germany). The test performance of the commercial antibody assay was previously verified in an in-house validation study using sera from negative and positive controls.^18^ According to the manufacture’s product sheet, applying a cut-off of >1·1 for defining seropositivity results in an estimated sensitivity of 94·4% (at >10 days of infection) and specificity of 99·6%. Next, we performed two additional confirmatory tests in all those individuals whose ELISA assay results were either positive (i.e. >1·1) or borderline (i.e. between 0·8 and 1·1). Confirmatory tests consisted of an in-house recombinant immunofluorescent test and a plaque reduction neutralisation test (PRNT) to specifically check for the presence of neutralizing antibodies against SARS-CoV-2.^21^

### Questionnaires

Data on physical and mental health were collected through an extensive questionnaire addressing current demographic, living and socioeconomic conditions, co-morbidities, medication and substance use, as well as COVID-19 related symptoms. The questions were selected taking account of other ongoing and developing COVID-19 related studies (especially the various “COVID-19 Host Genetics Initiative” cohorts,^22^ particularly the Lifelines study^23^; https://www.covid19hg.org), to facilitate future data harmonization, sharing and collaboration.

### Statistical analysis

Descriptive statistics are presented as means and 95% confidence intervals for continuous variables or numbers and percentages for categorical variables. Generalized estimating equations (GEE) with an independent covariance structure within household units were used to account for potential correlations between members of the same household. We used GEE with a logistic link function to estimate seroprevalence. We also applied GEE with either a logistic or Gaussian link function to assess which factors were associated with seropositivity or IgG ratio, respectively, while adjusting for potential confounders. We specifically assessed whether age, sex, education (as a measure of socioeconomic status), pre-existing medical conditions, vaccination, body mass index (BMI), smoking or alcohol consumption were associated with these outcomes. In addition, we assessed the relation between the number of symptoms experienced since January 1^st^, 2020, until the time of blood withdrawal and seropositivity. The GEE confidence intervals were based on the robust Huber-White sandwich variance estimator. In case of subgroups with zero counts, Fisher’s exact test was used instead for intergroup comparisons. All analyses were performed in R (base version 3.6.1) and a two-tailed *p*-value of < 0·05 was considered statistically significant.

## RESULTS

### Group I

#### Prevalence estimates

Cohort characteristics and the serosurvey results of Group I are displayed in **Table 1** and **Figure 1**. The participants originated from a total of 3983 different households, including 778 households with two and five households with three participants. Sixteen of the 46 individuals with a positive ELISA test result had neutralizing antibodies, whereas this was the case for only one of the 36 persons with an ELISA result within the borderline range (**Figure 1**). The estimated seroprevalence was 0·97% (95% CI: 0·72-1·30) by ELISA assay and 0·36% (95% CI: 0·21-0·61) by PRNT. The seroprevalence estimates were neither associated with age (OR 1·00 (95% CI: 0·98-1·02) for ELISA, and OR 0·98 (95% CI: 0·94-1·03) for PRNT) nor sex (male vs. female OR 0·94 (95% CI: 0·54-1·66) for ELISA, and 0·56 (95% CI: 0·23-1·40) for PRNT).

**Table 1.**
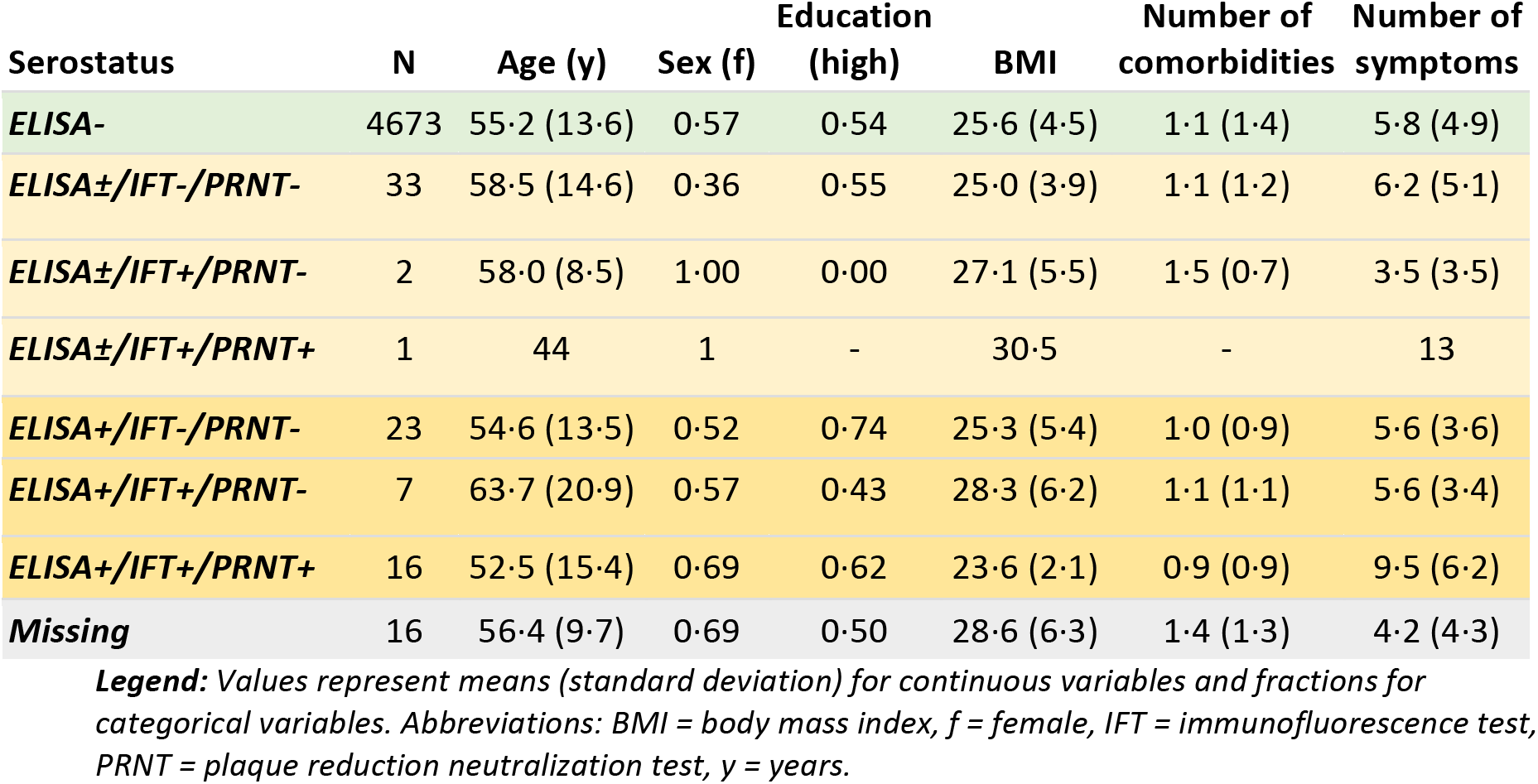
Sample characteristics (*Group I*) stratified by serostatus.

| Serostatus | N | Age (y) | Sex (f) | Education (high) | BMI | Number of comorbidities | Number of symptoms |
| --- | --- | --- | --- | --- | --- | --- | --- |
| **ELISA-** | 4673 | 55.2 (13.6) | 0.57 | 0.54 | 25.6 (4.5) | 1.1 (1.4) | 5.8 (4.9) |
| **ELISA±/IFT-/PRNT-** | 33 | 58.5 (14.6) | 0.36 | 0.55 | 25.0 (3.9) | 1.1 (1.2) | 6.2 (5.1) |
| **ELISA±/IFT+/PRNT-** | 2 | 58.0 (8.5) | 1.00 | 0.00 | 27.1 (5.5) | 1.5 (0.7) | 3.5 (3.5) |
| **ELISA±/IFT+/PRNT+** | 1 | 44 | 1 | - | 30.5 | - | 13 |
| **ELISA+/IFT-/PRNT-** | 23 | 54.6 (13.5) | 0.52 | 0.74 | 25.3 (5.4) | 1.0 (0.9) | 5.6 (3.6) |
| **ELISA+/IFT+/PRNT-** | 7 | 63.7 (20.9) | 0.57 | 0.43 | 28.3 (6.2) | 1.1 (1.1) | 5.6 (3.4) |
| **ELISA+/IFT+/PRNT+** | 16 | 52.5 (15.4) | 0.69 | 0.62 | 23.6 (2.1) | 0.9 (0.9) | 9.5 (6.2) |
| **Missing** | 16 | 56.4 (9.7) | 0.69 | 0.50 | 28.6 (6.3) | 1.4 (1.3) | 4.2 (4.3) |

**Legend:** Values represent means (standard deviation) for continuous variables and fractions for categorical variables. Abbreviations: BMI = body mass index, f = female, IFT = immunofluorescence test, PRNT = plaque reduction neutralization test, y = years.

**Figure 1:**
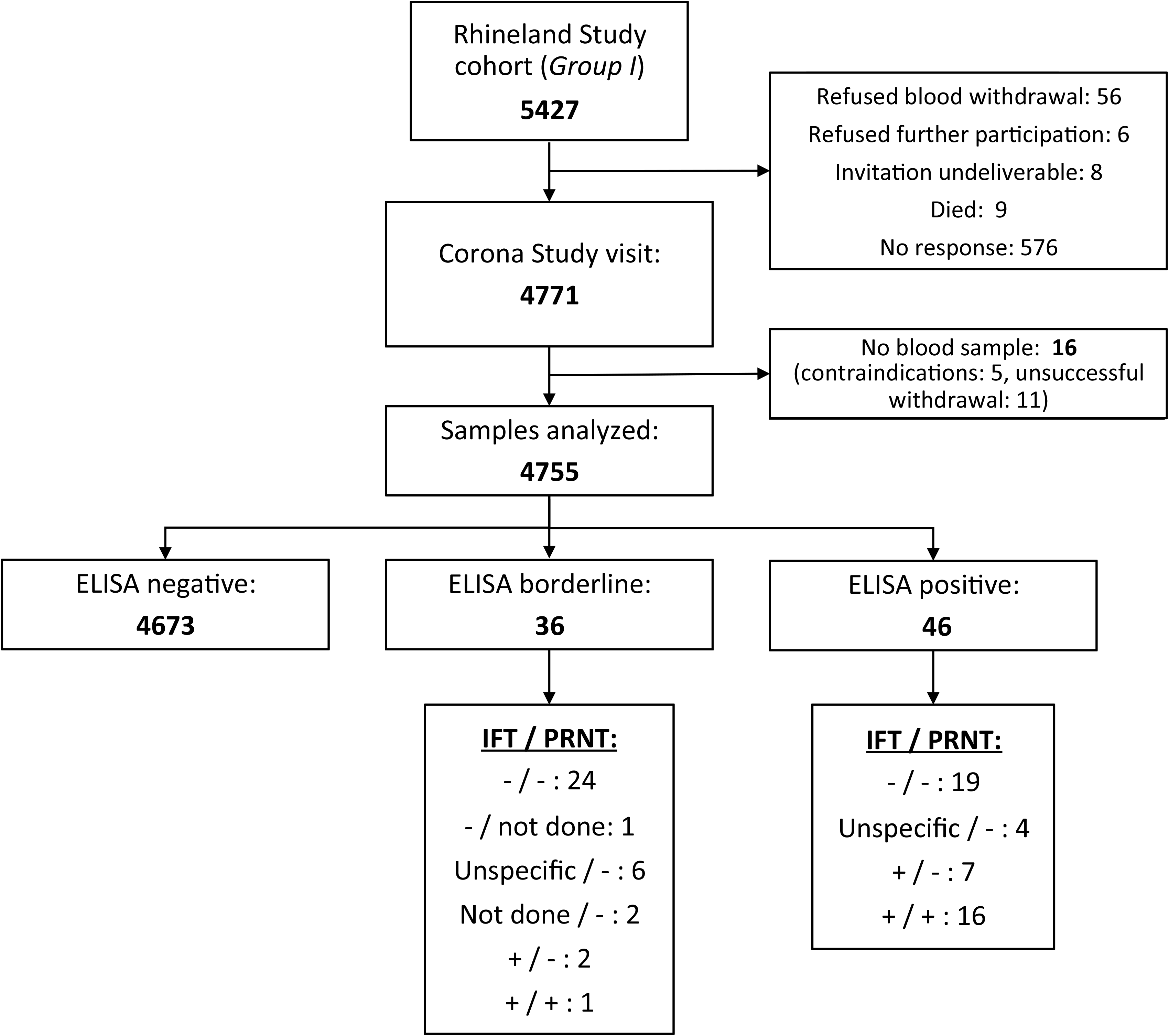
Flow chart. Overview of the number of participants, their test results as well as reasons for non-participation or missingness. A minus or a plus sign indicates a negative or positive confirmatory test result, respectively. Abbreviations: IFT = immunofluorescence test, PRNT = plaque reduction neutralization test.

#### Factors associated with the presence of neutralizing antibodies

The 17 individuals with neutralizing antibodies came from 15 different households, including two households with two cases each and two households with one participant with and one without neutralizing antibodies. Only one individual had suffered from severe COVID-19 requiring hospitalization and intensive care treatment. The other 16 individuals had not required hospital care and were therefore assumed to have had asymptomatic or mild to moderate infection: All of them reported having experienced at least one symptom since January 1^st^, 2020 (**Supplementary Figure 1**), with the odds of having neutralizing antibodies increasing with 12% (OR 1·12, 95% CI: 1·04 – 1·21) for each additional symptom reported. Apart from living with a person with a confirmed or suspected SARS-CoV-2 infection and the number of experienced symptoms, other factors – including education, body mass index, comorbidity, alcohol consumption, smoking and vaccination against seasonal influenza, pneumococcus or tuberculosis – were not associated with the presence of neutralizing antibodies (**Figure 2A**); whereas a reduced sense of taste or smell, fever in the last month, chills or hot flashes, pain while breathing, pain in the arms or legs, as well as muscle pain and weakness were all significantly associated with the presence of neutralizing antibodies (ORs ranging from 3·44 to 9·97, all p < 0·018; **Figure 2B**). Neither the total number of comorbidities nor the presence of a particular comorbidity was associated with the presence of neutralizing antibodies (**Figure 2A** and **Supplementary Figure 2**).

**Figure 2.**
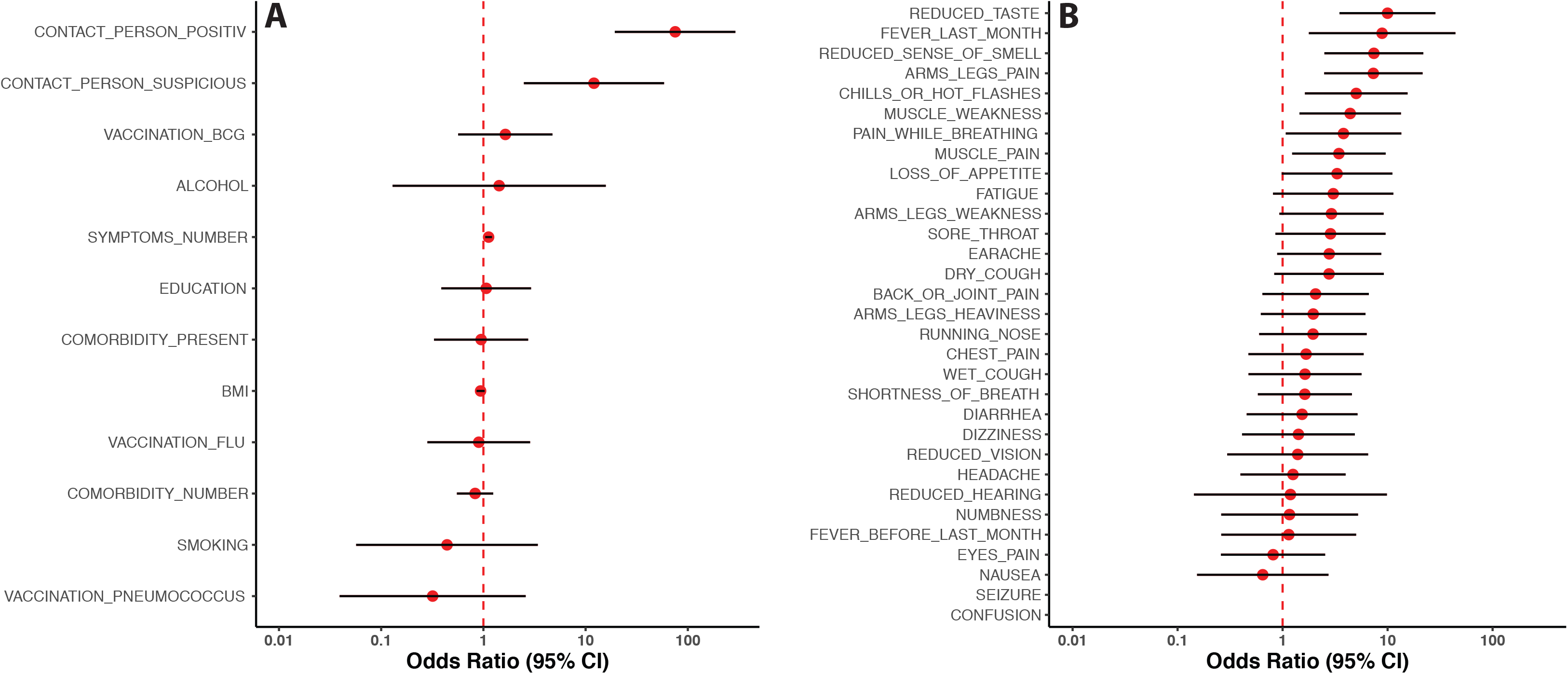
Factors associated with the presence of neutralizing antibodies. Living in the same household with a person with confirmed or suspected SARS-CoV-2 infection as well as a higher number of reported symptoms were significantly associated with the odds of having neutralizing antibodies (**A**). A reduced sense of taste or smell, fever in the last month, pain in arm/legs, chills/hot flashes, pain while breathing as well as muscle weakness and pain were significantly more often reported by individuals with versus those without neutralizing antibodies (**B**); seizures and confusion were not reported in the seropositive group, and because of a very low background prevalence, the associated odds ratios could not be estimated reliably. All estimates are adjusted for age, sex and household clustering. The odds ratio point estimates and the corresponding 95% confidence intervals are depicted by the red dots and whiskers, respectively, using a logarithmic scale.

#### Factors associated with the presence of neutralizing antibodies in ELISA+ individuals

In the subgroup of 46 ELISA+ individuals, those with neutralizing antibodies had a significantly higher antibody response as compared to those without neutralizing antibodies (age- and sex-adjusted difference in IgG ratio: 2·62, 95% CI: 1·81 – 3·43) (**Figure 3A**). In addition, only in those with neutralizing antibodies the IgG response significantly increased with age (0·08 per year (95% CI: 0·05 – 0·12) in the ELISA+/PRNT+ subgroup, and 0·05 per year (95% CI: −0·002 – 0·10) in the ELISA+/PRNT-subgroup; **Figure 3B**).

**Figure 3.**
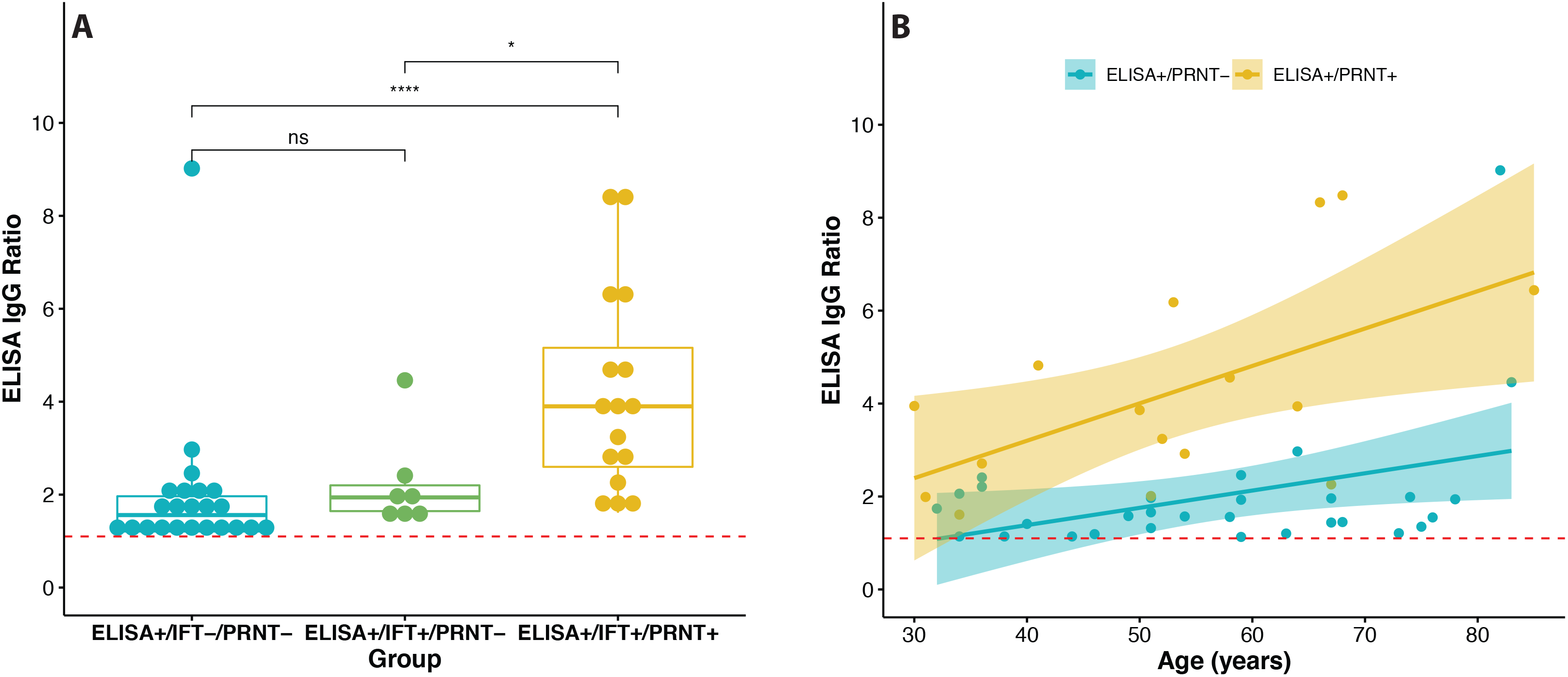
Relation between IgG response and neutralizing antibodies. Individuals with neutralizing antibodies had a significantly higher IgG antibody response as represented by the ELISA IgG ratio (**A**, a minus or a plus sign indicates a negative or positive test result, respectively). Only in the ELISA+/PRNT+ subgroup there was a significantly higher IgG response with increasing age (**B**). The red dotted lines indicate the threshold for a positive ELISA test result. Please refer to the main text for further details. Abbreviations: IFT = immunofluorescence test, PRNT = plaque reduction neutralization test; ns = not significant, * p < 0.05 and **** p < 0.0001 by the non-parametric Wilcoxon test.

None of the 30 ELISA+/PRNT-individuals reported living in a household with a member with a previously confirmed SARS-CoV-2 infection, whereas three ELISA+/PRNT+ individuals indicated living together with a person with a previously confirmed SARS-CoV-2 infection (Fisher’s exact test p = 0·05). Neither age (OR 0·98, 95% CI: 0·94 – 1·02) nor sex (male *vs*. female OR 0·47, 95% CI: 0·14 – 1·6) differentiated between the two groups. Those with neutralizing antibodies reported having experienced more symptoms (OR 1·19, 95% CI: 1·02 – 1·38), whereas other factors – including education, body mass index, comorbidity, alcohol consumption, smoking and vaccination against seasonal influenza, pneumococcus or tuberculosis – did not differentiate between the two groups (**Figure 4A**). Fever (in the last month) and earache were only reported in the group with neutralizing antibodies by two and four individuals, respectively (Fisher’s exact test p-values of 0·14 and 0·02 for fever and earache, respectively). The odds of neutralizing antibody seropositivity significantly increased with loss of appetite, muscle weakness, chills or hot flashes and a reduced sense of taste (**Figure 4B**).

**Figure 4.**
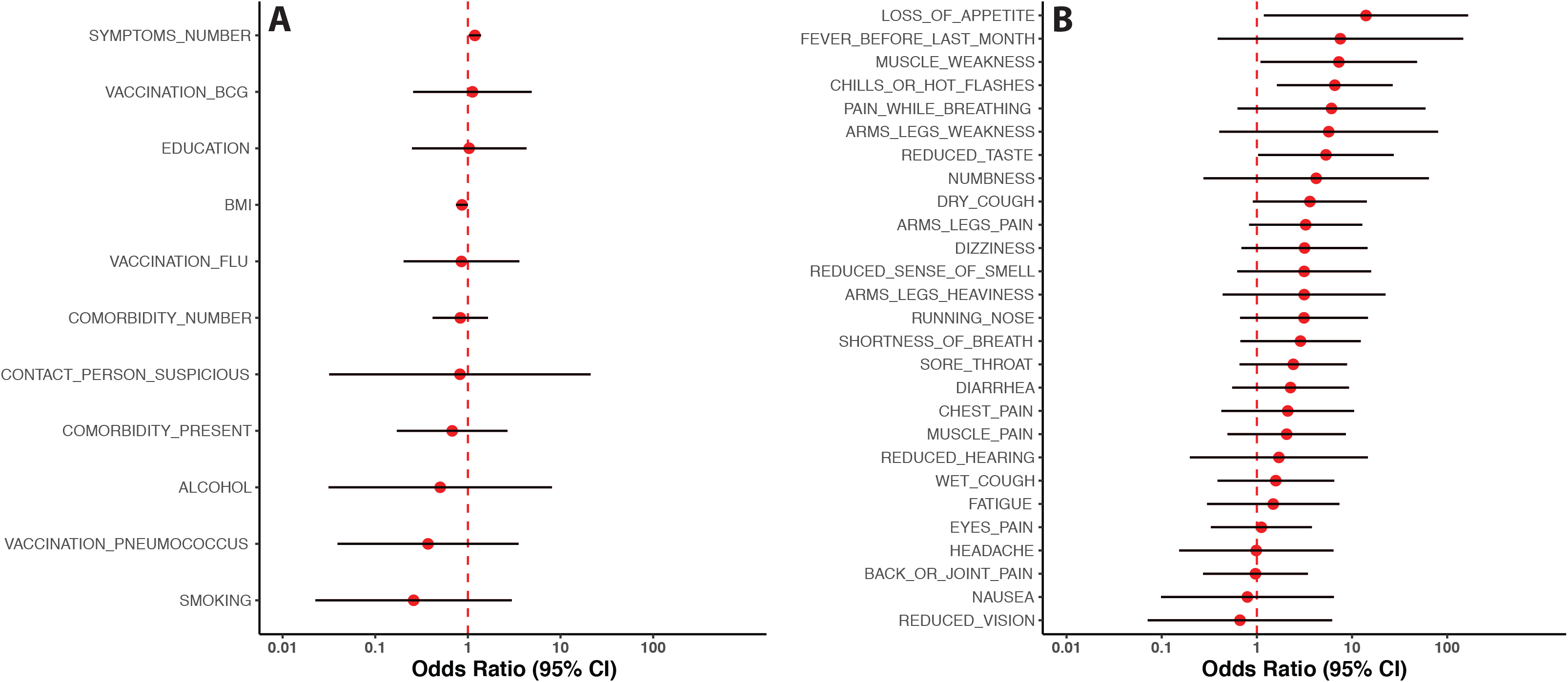
Factors differentiating between ELISA+ individuals with and without neutralizing antibodies. A higher number of reported symptoms was significantly associated with a higher odds of having neutralizing antibodies (**A**). Loss of appetite, muscle weakness, chills or hot flashes as well as a reduced sense of taste were significantly more often reported by individuals with versus those without neutralizing antibodies (**B**). All estimates are adjusted for age, sex and household clustering. The odds ratio point estimates and the corresponding 95% confidence intervals are depicted by the red dots and whiskers, respectively, using a logarithmic scale.

#### Follow up of people with borderline ELISA results

Twenty-seven of the 36 individuals in Group I with a borderline ELISA test returned for follow-up testing after a median of 28 days (range 20 to 41 days). At follow-up the IgG ratio had increased beyond 1·1 in six of these individuals; however, neutralizing antibodies could not be detected in any participant at the follow-up visit, not even in the individual in whom the presence of neutralizing antibodies was confirmed at the baseline visit (**Figure 5**).

**Figure 5.**
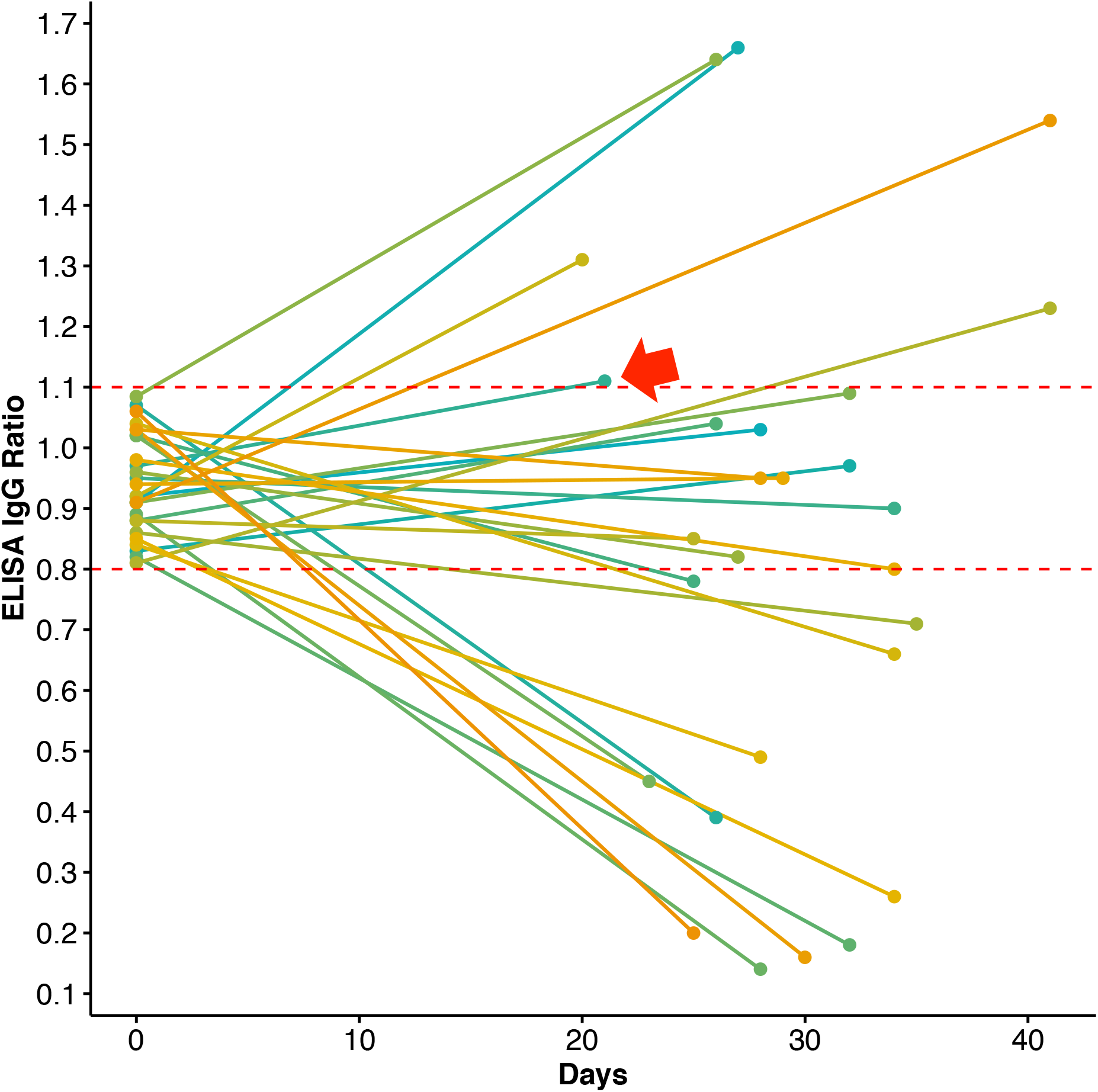
Time course of the antibody response in individuals with an indeterminate ELISA test result. All individuals with an IgG ratio in the indeterminate range (i.e. between 0.8 and 1.1) at baseline were reassessed after at least 20 days. The arrow marks the individual who had detectable neutralizing antibodies at baseline, but not at the follow-up visit. Neutralizing antibodies were not detectable in any participant at follow-up. The horizontal dotted lines represent the borders of the indeterminate range.

### Group II

A summary of the sample characteristics and test results of this group is presented in **Supplementary Table 1**. The seroprevalence in Group II was 1·94% (95% CI: 0·84–4·42) by the ELISA assay and 1·39% (95% CI: 0·49–3·85) by PRNT. Compared to Group I, the odds of a positive ELISA test result were two-fold higher in Group II, although the OR did not reach statistical significance (OR 2·03, 95% CI: 0·82–4·99). The odds of having neutralizing antibodies were almost four-fold higher in Group II compared to Group I (OR 3·88, 95% CI: 1·20–12·55). Groups I and II did not differ with respect to age, sex and number of comorbidities (all p ≥ 0·07).

However, a higher number of reported symptoms was associated with a slightly lower odds of being in Group II (OR 0·97, 95% CI: 0·95 – 1·00). Importantly, however, the proportion of people who reported having a household member with a confirmed SARS-CoV-2 infection was substantially higher in Group II compared to Group I (1·67% vs. 0·36%, adjusted p = 0·007). In addition, a higher proportion of people in Group II, as compared to Group I, reported having been previously tested positive for a SARS-CoV-2 infection (1·11% vs. 0·23%, adjusted p = 0·016).

## DISCUSSION

We present the findings of the first wave of the largest population-based seroepidemiological cohort study to date in Germany. We found an extremely low prevalence of SARS-CoV-2 seropositivity in Bonn, a middle-large city in the western part of Germany with a relatively low burden of COVID-19, both regionally and internationally.^16,17^ In addition, we found that: *1)* only about one third of the individuals who tested positive on a widely used quantitative immunoassay had detectable levels of serum neutralizing antibodies; *2)* both the magnitude of the antibody response, as reflected by the IgG ratio, the total number of symptoms experienced, as well as the presence of particular symptoms were associated with the presence of neutralizing antibodies in those with a positive immunoassay test result; *3)* in those with a borderline immunoassay result the presence of neutralizing antibodies was extremely rare, and -in the only confirmed case-transient, and *4)* self-referral bias can lead to substantial overestimation of seroprevalence.

As of June 30^th^, there were a total of 759 confirmed SARS-CoV-2 infections (including three COVID-19 related deaths) in Bonn, a city with about 330,000 inhabitants, yielding a prevalence of 0·23%.^24^ Our seroprevalence estimates are 1·6 to 4·2 times higher (based on the presence of neutralizing antibodies or a positive ELISA test result, respectively). The study sample did not include people younger than 30 years old and is, therefore, not representative of the entire Bonn population. Nevertheless, our findings suggest that a considerable number of individuals who had been infected have evaded case ascertainment by the local health regulatory agencies. Our findings also confirm that limited exposure of the local population to SARS-CoV-2 most likely accounts for the relatively low rates of regional COVID-19-related hospitalizations and mortality, supporting the efficacy of early implementation of social distancing and confinement measures in Germany.^16,17^ The current seroprevalence estimates are lower than the two other German community-based serosurveys.^14,15^ However, both these previous serosurveys were conducted in communities following super-spreading events, and are unlikely to reflect the state of other regions in Germany with relatively low COVID-19 burden.^14,15^

Neutralizing antibodies were detected in only about one third of the participants who tested positive on a widely used immunoassay. At least two explanations may account for this finding. First, the individuals who tested positive on the immunoassay but not on PRNT, may be false positives, e.g. due to cross-reactivity with antibodies against other coronaviruses. Based on the reported specificity of the ELISA assay that we used and assuming a zero prevalence, we would have expected 19 false positives.^18^ Second, this group may also include individuals who were infected by SARS-CoV-2, but who either did not develop neutralizing antibodies or lost them in the period following infection. Indeed, we could not detect neutralizing antibodies in 6 out of 15 people who reported to have had a SARS-CoV-2 infection in the past. This latter finding is also in line with recent reports indicating that neutralizing antibodies may not develop in asymptomatic or only mildly symptomatic individuals and, especially in this group, may wane relatively quickly in the period following infection.^19^ Given that neutralizing antibodies are thought to be a major component of adaptive immunity and their levels also strongly correlate with the number of SARS-CoV-2 specific T-cells,^25^ these findings thus suggest that even after infection a large proportion of individuals in the general population may become susceptible again to SARS-CoV-2 infection, making “herd immunity” even more difficult to achieve.

Among persons with a positive immunoassay result, the magnitude of the IgG response, the number of previously experienced symptoms, as well as the prior occurrence of particular symptoms – including loss of appetite, muscle weakness, chills or hot flushes and reduced taste – were strongly associated with the probability of having neutralizing antibodies. This suggests that predictive models can be developed, based on a combination of clinical characteristics and the magnitude of the antibody response to estimate the probability of a person having neutralizing antibodies. Testing for neutralizing antibodies is currently very labor intensive and can only be reliably performed in specialized laboratories. Good prediction models could be useful to better estimate the actual population immunity level in regions without access to such advanced testing facilities.

Self-referral or volunteer bias could inflate seroprevalence estimates.^26^ In order to estimate the magnitude of this effect, we also sampled a group of spontaneous volunteers from the same region who were not part of the original Rhineland Study cohort, but who expressed interest in the serosurvey. After formal invitation of these individuals, the response rate was almost 30% lower compared to the original cohort of participants, whereas the seroprevalence estimates were two to four-fold higher (based on the presence of neutralizing antibodies or a positive ELISA test result, respectively). It appeared that the main reasons for self-selection were not so much the presence of symptoms, but a previously confirmed SARS-CoV-2 infection or the presence of a close contact with a previous SARS-CoV-2 infection. These findings thus illustrate the profound impact of selection-bias on seroprevalence estimates – likely a major source for the large heterogeneity of the findings of many previous serosurveys – and thereby underscore the critical importance of cohort-based analyses that allow for accurate quantification of response rate and reasons for (non-)response.

Our study has both strengths and limitations. By implementing this serosurvey in an ongoing community-based prospective cohort study, we were able to quickly reach and mobilize a large group of participants and achieve a very high response rate, thereby minimizing the risk of selection bias. This approach will also ensure systematic follow-up in the future, including serological reassessments, in a cohort of already deeply phenotyped individuals, allowing for the tracking of the evolution of the pandemic, identification of the determinants of susceptibility and resilience to SARS-CoV-2 infection, as well as elucidation of any long-term physical and mental health sequelae. The limitations of our study include the lack of a representative sample of the general population, since our study only enrolled individuals ≥ 30 years, and the fact that we did not assess SARS-CoV-2 specific T-cell response, another critical component of acquired immunity.^27,28^ Nevertheless, given the low (neutralizing antibody) seroprevalence, it is highly unlikely that the population seroprevalence in this region would materially differ from our estimates. Moreover, we are already planning to include indices of cellular immunity in follow-up serosurveys.

In conclusion, we found a relatively low SARS-CoV-2 seroprevalence in participants of a large-scale community-based cohort study in Bonn, Germany (until June 30^th^, 2020), with neutralizing antibodies detectable in only one third of those with a positive immunoassay result. These findings indicate that almost the entire population in this region remains susceptible to SARS-CoV-2 infection, and thus warrant continued vigilance by the health authorities.

## Data Availability

The Rhineland Study's dataset is not publicly available because of data protection regulations. Access to data can be provided to scientists in accordance with the Rhineland Study's Data Use and Access Policy. Requests for further information or to access the Rhineland Study's dataset should be directed to.

## AUTHOR CONTRIBUTIONS

NAA, VMC, AS, CD, and MMBB conceived the study. NAA, VMC, FMdV, AKCE, THS, AS, CD, and MMBB participated in the study design. FMdV, AKCE, THS, and AS coordinated the data and biosample collection and processing. VMC, MLS, AR, and CD supervised and coordinated the performance of the serological assays. NAA did the data analyses. NAA and MMBB drafted the first version of the manuscript. All authors contributed to data interpretation, participated in revising the manuscript for important intellectual content, and read and approved the final manuscript.

## ACKNOWLEDGEMENTS

The authors would like to thank all the staff members and participants of the Rhineland Study. We would especially like to thank Theda Backen, Nüket Cebi, Karin Heubach, Angelika Klein, Stefan Kurth and Ina Plettner for their dedicated support in the planning and management of this research project; Nersi Alaeddin, Felix Ernst, Elvire Landstra, Dan Liu, Marina Santos, Valentina Talevi, and Weiyi Zeng for valuable assistance with the processing of the large number of bio-samples; and Simon Harmata and André Medek for relentless IT-support.

## Supplementary Material

**Supplementary Table 1.** Sample characteristics *(Group II)* stratified by serostatus.

| Serostatus | N | Age (y) | Sex (f) | Number of comorbidities | Number of symptoms |
| --- | --- | --- | --- | --- | --- |
| ELISA- | 350 | 55.1 (12.5) | 0.61 | 1.0 (1.3) | 5.2 (4.6) |
| ELISA±/IFT-/PRNT- | 3 | 52.3 (11.0) | 0.33 | 2.7 (2.3) | 3.3 (3.5) |
| ELISA+/IFT+/PRNT- | 2 | 68.0 (17.0) | 0.50 | 0.5 (0.7) | 6.5 (2.1) |
| ELISA+/IFT+/PRNT+ | 4 | 55.2 (4.6) | 0.75 | 1.0 (1.2) | 8.0 (6.2) |
| ELISA+/IFT±/PRNT+ | 1 | 58 | 0 | 1 | 3 |

**Legend:** Values represent mean (standard deviation) for continuous variables and fractions for categorical variables. Abbreviations: f = female, IFT = immunofluorescence test, PRNT = plaque reduction neutralization test, y = years.

**Supplementary Figure 1.**
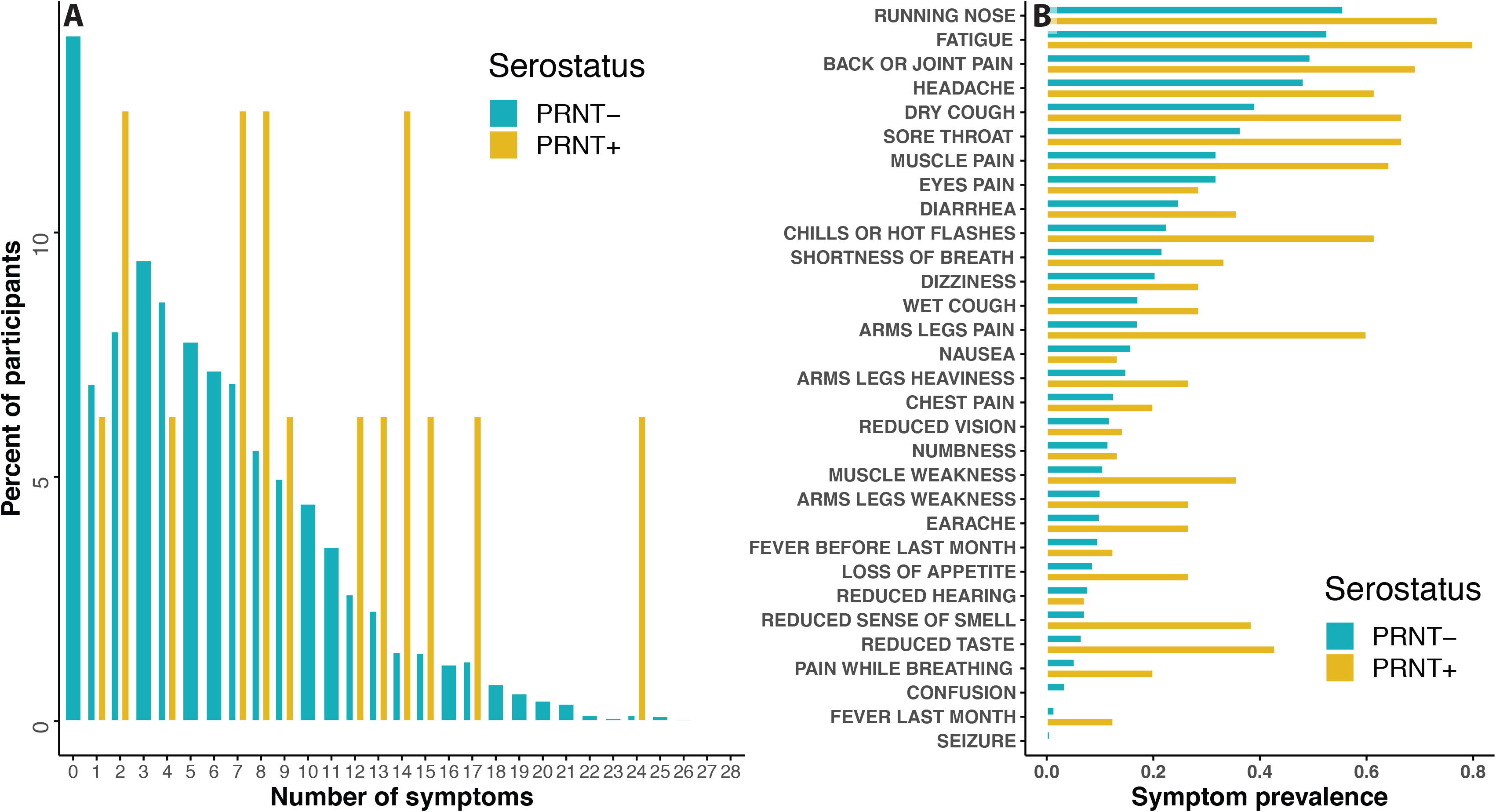
Proportion of participants reporting symptoms, stratified by serostatus and number of symptoms (**A**) or type of symptoms (**B**). Abbreviations: PRNT = plaque reduction neutralization test.

**Supplementary Figure 2.**
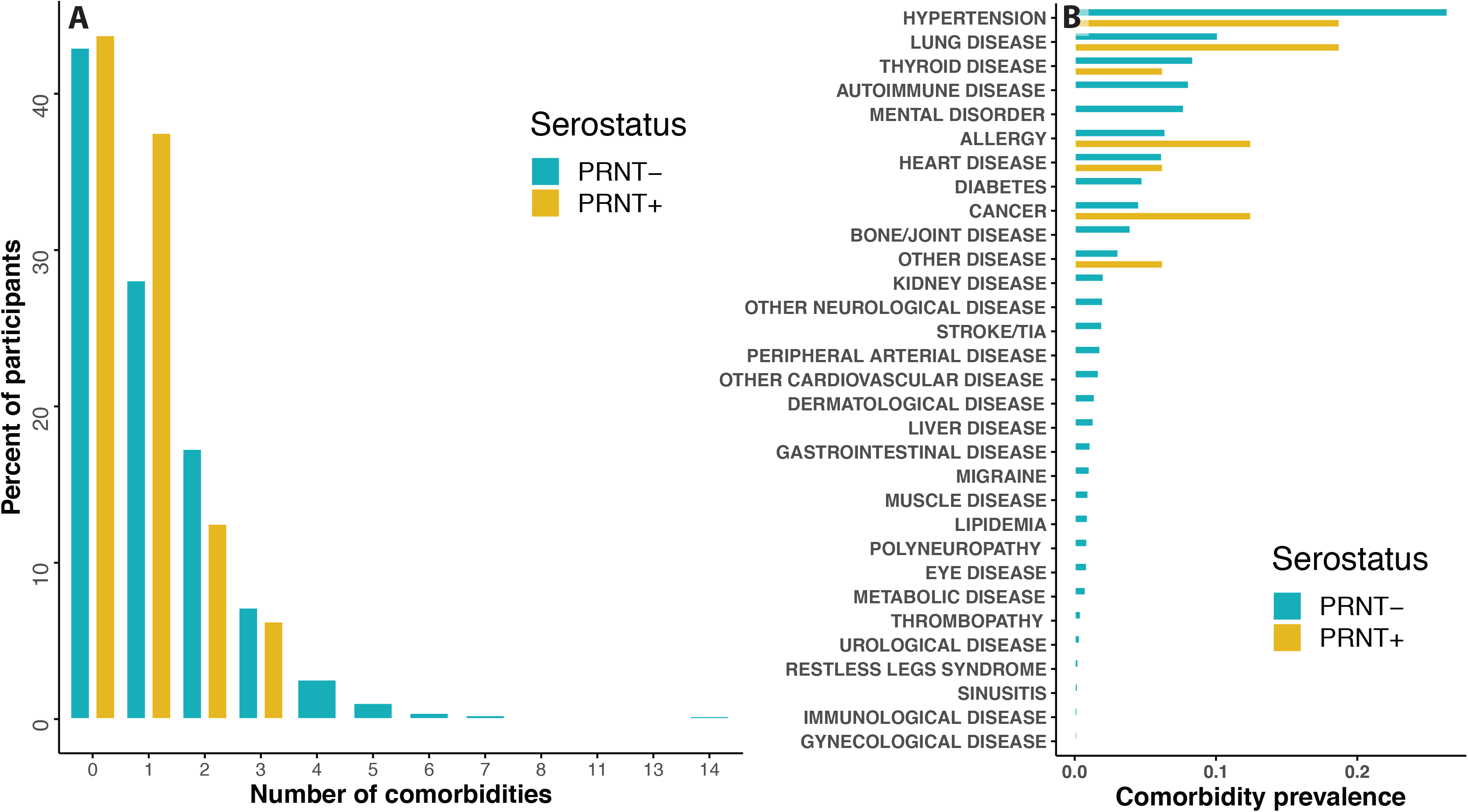
Proportion of participants reporting comorbidities, stratified by serostatus and number of comorbidities (**A**) or type of comorbidity (**B**). Abbreviations: PRNT = plaque reduction neutralization test.

